# Simple Mathematics on Covid-19 Expansion

**DOI:** 10.1101/2020.03.17.20037663

**Authors:** F. Javier García de Abajo

## Abstract

We review the Kermack and McKendrick model of epidemics and apply it to Covid-19. Despite the simplicity of this model, solid conclusions are extracted that can assist potential decisions on the strategy to combat the outbreak, essentially configuring a scenario ranging from short-term suppression to long-term mitigation depending on the achieved reduction in the contact number.

---

In their so called SIR model, Kermack and McKendrick [1] formulated a rate equation describing the expansion of infections in the population under the assumption of a uniform transmission rate and exposure of all individuals. In brief, they examined the fraction of susceptible people *s*(*t*) who have not been infected at a given time *t*, as well as the fraction of infected people *i*(*t*) that can still be contagious and the fraction of recovered people *r*(*t*) = 1 −*s*(*t*) − *i*(*t*) that have been previously infected but are no longer contagious because they are either cured or dead. The latter evolves in time according to [2]

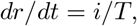

where *T* is a characteristic *lifetime* defining the 1*/e* decay in the probability of an individual to transit from infected (*i*) to recovered or dead (*r*). Also, the uninfected fraction is depleted according to

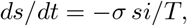

which describes an exponential transit from susceptible (*s*) to infected (*i*) at a rate that is *σ* times faster than the decay from infected to recovered. We can roughly understand *T* as the average time during which an infected individual remains contagious, while *σ* is the contact number, also known as transmission number (i.e., the number of individuals that become infected on average due to exposure to one previously infected specific individual). The expansion of Covid-19 presents a dynamical scenario and we expect it to remain so and to depend on local habits, implementation of social distancing, and response of the population in each specific region. This model can then be used to estimate the evolution of the outbreak at a local scale over a limited period of time in which *σ* remains approximately constant. Extrapolations to the global scale are more challenging and may require an extension of the model combined with the availability of reliable relevant parameters.

The above equations for the SIR model can be conveniently solved as a function of the normalized time *τ* = *t/T*, in terms of which we have

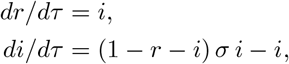

together with *s* = 1 −*r*− *i*. Initial conditions at the beginning of the outbreak (*t* = 0) are *r* = 0 and *i* ≪ 1. The actual value of *i* at *t* = 0 determines the time needed to reach a maximum in *i*, but not the magnitude of this maximum. Solutions of the above equations are widely available in the literature, including phase diagrams of epidemics for fixed *σ* [2]. We base the remainder of the present analysis of Covid-19 on the solutions shown in Figs. 1 and 2.

**FIG. 1:**
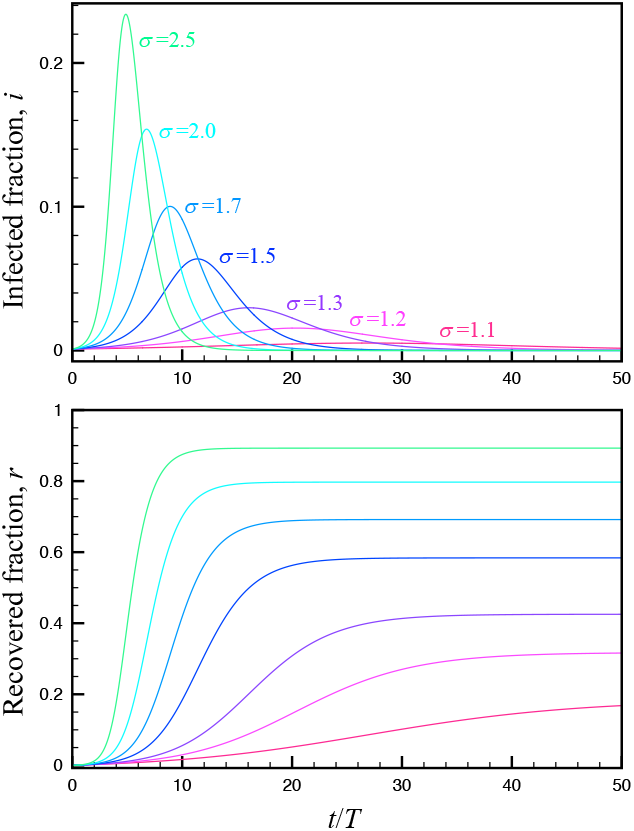
Solutions of the SIR model for different values of the contact number *σ*. We show the infected (top) and recovered (bottom, including deaths) fractions as a function of time *t* normalized to the active time *T* over which an infected person remains contagious. We take *r*(0) = 0 and *i*(0) = 10^−3^ at *t* = 0. The obtained curves are simply displaced to the right if smaller values of *i*(0) is assumed.

**FIG. 2:**
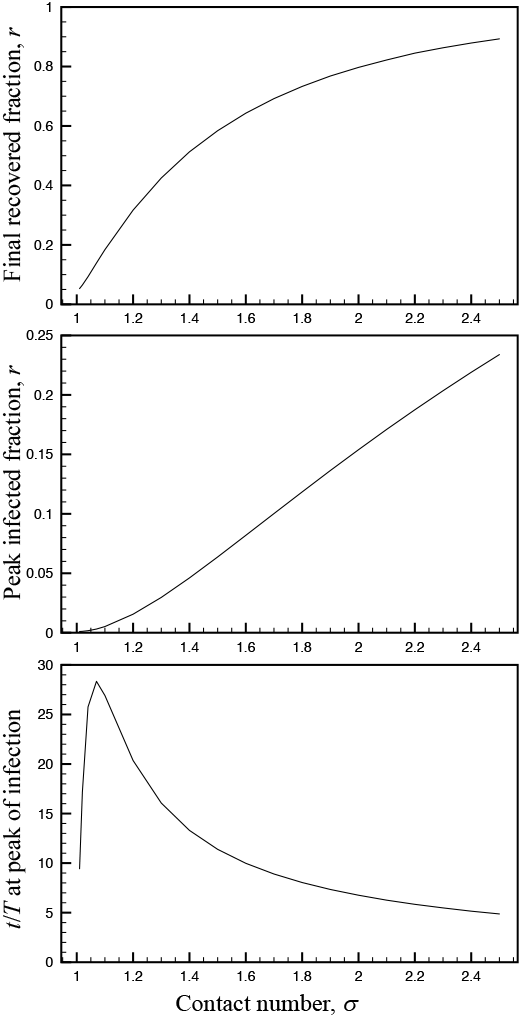
Evolution of the asymptotic fraction of recovered (top) and infected (middle) people as a function of contact number *σ*. The bottom plot shows the normalized time at which the peak of infection is reached for *r*(0) = 0 and *i*(0) = 10^−3^.

We are currently at the beginning of the outbreak, so the infected and recovered fractions *i* and *r* are still small. The evolution of the infection should then depend on the contact number *σ*, for which values in the 1.5-3 range have been estimated, although conclusive data are still lacking. This number might be affected by mutated variants of the virus, which should also require a more sophisticated analyses beyond the SIR model.

For an order-of-magnitude estimate of the impact of the outbreak, assuming a likely value of *σ* ∼ 2.5, the peak in infected fraction exceeds 0.2 (Fig. 1, top). This figure has to be considered together with the mortality of the infection, currently believed to be ∼ 1% with medical assistance for the most serious cases (∼ 10%) and *>* 3% without medical assistance. With a ≪ 1% fraction of hospital beds per inhabitant in most countries, and even lower capacity of the required type of help (e.g., oxygen concentrators and respiratory assistance) for the most serious cases of Covid-19 infection, the evolution with a maintained contact number *σ* ∼ 2.5 implies a 0.03× 0.89 ≈2.7% final death toll.

Social distancing can bring down the peak infected fraction, and consequently, also the overall impact of the infection, both because medical treatment can be provided to a larger fraction of the infected population and because herd immunity is reached with a lower recovered fraction (Fig. 1). Effectively, social distancing can be understood in the light of the SIR model as a reduction of the value of *σ*. Still, the simulations presented in Fig. 1 show that even with a reduction from *σ*∼ 2.5 to *σ*∼ 1.3 (a level similar to seasonal flu), the infected fraction stays well above hospital capacity, implying a 3% mortality rate of the infection, and eventually resulting in a 1.2% final death toll.

A pertinent question in this respect is whether the measures adopted by different countries can bring the effective value of *σ* down to a range sufficiently close to 1, or even below 1 (i.e., the condition for exponential attenuation of the outbreak), in order to reduce the above levels of final death toll. In this respect, the media have speculated that summer conditions could push *σ* below 1, a possibility that seems to be unlikely when examining the current evolution of the outbreak in the Southern hemisphere.

Here, we presented results normalized to the average time *T* during which an individual remains contagious. Currently, no conclusive values exist for this quantity, so for a rough estimate we can assume *T*∼ 2 −4 weeks. The fastest outcome of the pandemic (free evolution with *σ*∼ 2.5) then requires ∼ 20 − 40 weeks to reach a low fraction of infected people after maximum expansion of the virus (Fig. 1, top). With social distancing measures that bring *σ* down to a value reasonably close to 1, the process should take even longer, eventually extending over the period needed to develop a vaccine and produce a sufficient number of doses to reach herd immunity (ignoring the effect of eventual mutations). Additional information on the final fraction of the population that becomes infected, the peak fraction of infected people, and the time at which such peak occurs in units of *T* is provided in Fig. 2 as a function of the contact number *σ*. We should stress again the relevant question that arises, are the measures taken by different countries sufficient to produce a substantial departure down from the value of *σ* ∼ 2.5 that seems to rule the outbreak in the absence of social distancing? Are those measures enough to push *σ* close or below 1 in order to avoid a collapse of our medical systems?

Two models are being proposed at the moment: controlled spreading (UK, The Netherlands); and lockdown (China, Italy, Spain). While the former is a social experiment that goes beyond the scope of this short report, the latter could work if one approaches *σ* ≪1. To illustrate that, at the current early state (*i, r* ≪ 1), the evolution of the infected fraction can be approximated by a simplified version of the SIR model: *di/dτ* = − (1− *σ*) *i*. **In contrast to the long times required to deal with the situation by reducing** *σ*, **but staying above 1, the strategy of making** *σ <* 1 **leads to an exponential attenuation of** *i* ∼ e^−(1−*σ*)*τ*^ **with a characteristic decay time** *T/*(1 −*σ*). **This model thus supports the recommendation of making** *σ*≪1 **to deal with the problem in a time interval stretching a few times** *T* **(the Hubei solution)**.

## Data Availability

All data are contained in the write up of the paper.

## Notes

### Competing Interest Statement

The authors have declared no competing interest.

